# The cholinergic system and treatment response in subtypes of Alzheimer’s disease

**DOI:** 10.1101/2020.01.23.20018507

**Authors:** Alejandra Machado, Daniel Ferreira, Michel J. Grothe, Helga Eyjolfsdottir, Per M. Almqvist, Lena Cavallin, Göran Lind, Bengt Linderoth, Åke Seiger, Stefan Teipel, Lars U. Wahlberg, Lars-Olof Wahlund, Eric Westman, Maria Eriksdotter, for the Alzheimer’s Disease Neuroimaging Initiative

**Author notes:** Corresponding author at: Division of Clinical Geriatrics, Centre for Alzheimer Research, Department of Neurobiology, Care Sciences, and Society, Karolinska Institutet, NEO, Floor 7th, Blickagången 16, 141 52 Huddinge, Sweden., E-mail address (D. Ferreira). Shared first authorship (Alejandra Machado, Daniel Ferreira). Shared senior authorship (Eric Westman, Maria Eriksdotter). Data used in preparation of this article were obtained from the Alzheimer’s Disease Neuroimaging Initiative (ADNI) database (adni.loni.usc.edu/). As such, the investigators within the ADNI contributed to the design and implementation of ADNI and/or provided data but did not participate in analysis or writing of this report. A complete listing of ADNI investigators can be found at: http://adni.loni.usc.edu/wp-content/uploads/how_to_apply/ADNI_Acknowledgement_List.pdf.

## Abstract

**BACKGROUND:** The heterogeneity within Alzheimer’s disease (AD) seriously challenges the development of disease modifying treatments. We investigated volume of the basal forebrain, hippocampus, and precuneus in atrophy subtypes of AD, and explored the relevance of subtype stratification in a clinical trial on encapsulated cell biodelivery (ECB) of nerve growth factor (NGF) to the basal forebrain.

**METHODS:** Structural MRI data was collected for 90 amyloid-positive patients and 69 amyloid-negative healthy controls at baseline, 6-, 12-, and 24-month follow-up. The effect of the NGF treatment was investigated in 10 biopsy verified AD patients with structural MRI data at baseline and at 6- or 12-months follow-up. Patients were classified as typical, limbic-predominant, hippocampal-sparing, or minimal atrophy AD, using a validated visual assessment method. Volumetric analyses were performed using a region-of-interest approach.

**RESULTS:** All AD subtypes showed reduced basal forebrain volume as compared with controls. Limbic-predominant subtype showed fastest basal forebrain atrophy rate, whereas minimal atrophy subtype did not show significant volume decline over time. Atrophy rates of hippocampus and precuneus also differed across subtypes. The NGF treatment seemed to slow the rate of atrophy in precuneus and hippocampus, particularly in the hippocampal-sparing AD subtype.

**CONCLUSIONS:** The cholinergic system is differentially affected in distinct atrophy subtypes of AD, possibly contributing to their differential response to cholinergic treatment. Our findings suggest that future clinical trials should target specific subtypes of AD, or at least report treatment effects stratifying by subtype.

**Trial registration:** ClinicalTrials.gov identifier: NCT01163825. Registered 14 July 2010 - https://clinicaltrials.gov/ct2/show/NCT01163825

## 1. Background

Finding a cure for Alzheimer’s disease (AD) continues to be a major challenge. More than 200 AD clinical trials have failed to date [1], possibly due to the recruitment of heterogeneous populations. Different biological subtypes can be found in AD. Murray et al., (2011) showed that AD patients often have balanced neurofibrillary tangle (NFT) counts in the hippocampus and association cortex, i.e. the typical AD subtype. However, two other subtypes were also identified, corresponding to limbic-predominant and hippocampus-sparing AD, with NFT counts predominantly in the hippocampus or the association cortex, respectively. Structural magnetic resonance imaging (sMRI) can reliably track these subtypes *in vivo* [3], and has consistently identified a fourth subtype with minimal atrophy, i.e. the minimal atrophy AD subtype [4–8].

Currently approved treatments for AD are symptomatic and the most widely established treatments are cholinesterase inhibitors (ChEI) targeting the cholinergic system [1]. However, ChEI have limited effectiveness and alternative treatments targeting the cholinergic system are being investigated [9,10]. The basal forebrain is the major source of cholinergic innervation in the brain targeting the hippocampus and cortical areas [11,12]. AD patients with less hippocampal atrophy seem to respond better to ChEI [13]. This raises the hypothesis of whether hippocampal-sparing and minimal atrophy AD could have a better response to cholinergic treatment. Interestingly, hippocampal-sparing and minimal atrophy are the most frequent subtypes among patients with dementia with Lewy bodies (DLB) [14], who often respond well to ChEI [15]. Thus, impaired cholinergic system but relatively intact hippocampal function may be prognostic factors for a good response to ChEI [16]. However, no previous studies have investigated cholinergic system integrity or cholinergic treatment response across subtypes of AD.

We investigated impairment of the cholinergic system by analyzing atrophy in the basal forebrain and its target regions across the four subtypes of AD, both cross-sectionally and longitudinally. We then explored the effect of AD subtype on regional atrophy rates in AD patients with and without a cholinergic treatment consisting of encapsulated cell biodelivery (ECB) of nerve growth factor (NGF) to the basal forebrain. The atrophy rates of the treated sample (NGF cohort) where compared to the “expected” atrophy rates from an independent and untreated AD sample. This NGF treatment was an add-on to ChEI treatment since all patients were already under ChEI treatment. Targeted delivery of NGF has emerged as a potential therapy based on its regenerative effects on the basal forebrain cholinergic neurons [17–19]. Our AD patients treated with NGF are part of a study of targeted delivery of NGF to the basal forebrain over 6 or 12 months [20,21]. Hence, the present study includes a unique “experimental manipulation” of the basal forebrain in AD subtypes. We hypothesized that (1) the four AD subtypes would have significantly less volume in the basal forebrain at baseline compared with healthy controls; (2) the AD subtypes would show different baseline and atrophy rates of the basal forebrain with typical and limbic-predominant AD undergoing faster atrophy; (3) the different AD subtypes would show distinct correlations between longitudinal atrophy of the basal forebrain and longitudinal atrophy of the target regions; (4) the NGF treatment may have better response in patients with no hippocampal atrophy, i.e., slower atrophy rate than expected in hippocampal-sparing and minimal atrophy AD subtypes.

## 2. Method

### 2.1. Participants

A total of 90 AD patients and 69 healthy controls were selected from the ADNI-1 cohort [22]. All AD patients were amyloid β (Aβ)-positive and all healthy controls were Aβ-negative, using established cut-offs [23]. Participants’ selection and diagnostic criteria can be found on the ADNI webpage (http://www.adni-info.org). Stable doses of baseline medications, including ChEI (i.e., Aricept, Exelon or Reminyl), were permitted if listed in the ADNI procedures manual. The ADNI is a longitudinal multisite study from the United States and Canada launched in 2003 by the National Institute on Aging, the National Institute of Biomedical Imaging and Bioengineering, the Food and Drug Administration, private pharmaceutical companies and non-profit organizations (principal investigator Michael W. Wiener). The ADNI was approved by the institutional review board at each site. Informed consent was obtained from all participants.

In addition, 10 AD patients were recruited from the memory clinic at the Karolinska University Hospital (Huddinge, Sweden) (from here referred to as the NGF cohort). Inclusion criteria were: (1) a probable diagnosis of mild or moderate AD according to the NINCDS-ADRDA criteria [24], (2) aged 55–80 years, (3) a mini-mental state examination (MMSE) [25] score of 16–24, (4) living at home with a caregiver, and (5), stable treatment with similar ChEI as the ADNI cohort, for at least 9 months before enrollment, which remained stable during the study. All 10 AD patients underwent surgical implantation of NGF-releasing cell capsules, using encapsulated biodelivery bilaterally implanted into the basal forebrain. For details on study design, neurosurgical procedure and clinical follow-up, please see Wahlberg et al. 2012 and Eriksdotter et al. 2012. AD diagnosis was histopathologically confirmed in nine patients using cortical brain biopsies obtained during the surgical procedure [9]. In one patient, the biopsy failed and only provided fibrotic tissue. Diagnosis of the remaining patient was based on core clinical criteria and pathological CSF AD biomarkers. Exclusion criteria were the same as for the ADNI cohort, also including smoking.

### 2.2 MRI methods

#### 2.2.1. MRI data acquisition and processing

The MRI acquisition protocols were very similar for both cohorts and are described in Appendix A and elsewhere [26,27]. MRI data was collected at identical follow-up intervals in the ADNI and NGF cohorts, i.e. baseline, 6- and 12-month follow-ups. In addition, 24-month follow-up was also included for the ADNI cohort to investigate atrophy over a longer period (Table 1).

**Table 1.**
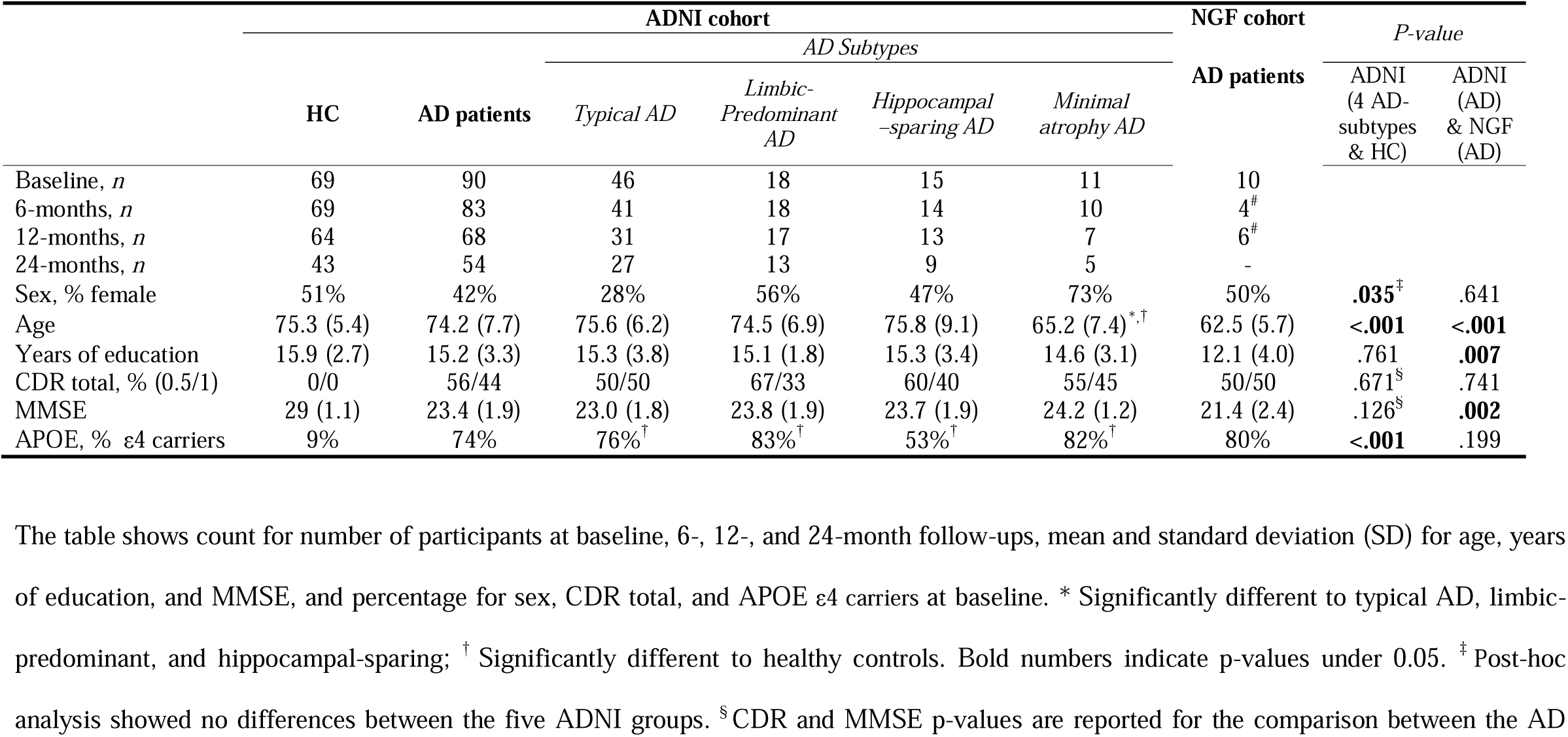
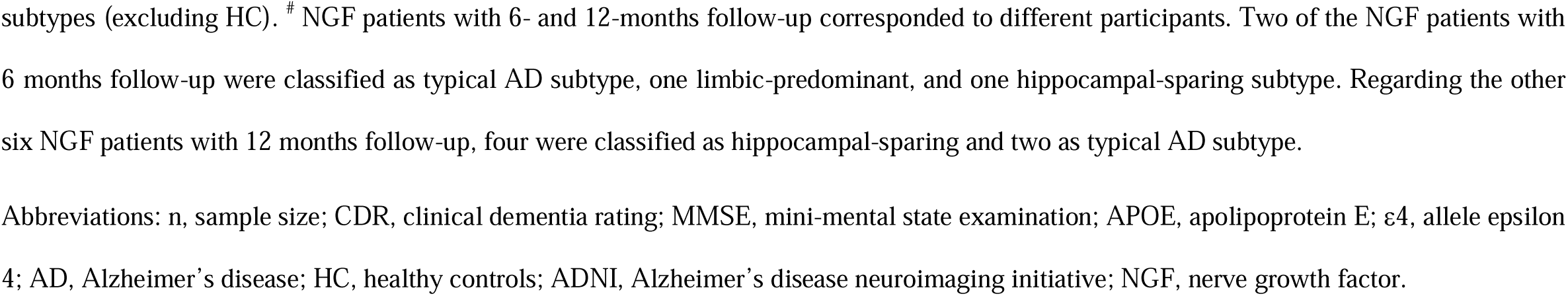
Baseline demographic and clinical characteristics of the ADNI and NGF cohorts.

The MRI data were processed using the statistical parametric mapping software (SPM8) and the voxel-based morphometry (VBM8) toolbox (http://dbm.neuro.uni-jena.de/vbm/). First, baseline and follow-up images of each individual were rigidly registered to each other and bias corrected for magnetic field inhomogeneities. Next, images were segmented into gray matter (GM), white matter (WM), and cerebrospinal fluid (CSF) partitions. GM and WM partitions from all subjects and time-points were then high-dimensionally registered to a customized template corresponding to the group’s anatomic mean using the DARTEL algorithm [28] (see Appendix A for more details). Flow fields resulting from this DARTEL registration were then used to warp the corresponding GM segments and voxel values were modulated to preserve the amount of GM volume present before warping.

#### 2.2.2. Regions of interest (ROIs)

The cholinergic space of the basal forebrain was defined using a stereotactic map of cholinergic basal forebrain nuclei in MNI standard space that was derived from combined post-mortem MRI and histologic staining as described in Kilimann et al. (2014). Other masks available in the SPM software were used to segment the precuneus (AAL atlas), the hippocampus and the primary somatosensory cortex (PSC) (anatomy toolbox) (Figure 1A). The hippocampus and precuneus are target regions of basal forebrain cholinergic projections [30]. The PSC was included as a negative control region [31]. Volumes from left and right hemispheres were summed up for the four masks. The masks defined in MNI space were warped to the DARTEL customized space and the GM volumes of the four ROIs were extracted for each individual and timepoint by summing up the modulated voxel values of the respective warped GM image. The total intracranial volume (TIV) was calculated as the sum of the total volumes of the GM, WM, and CSF partitions. ROI volumes were corrected for the TIV using residuals from linear regression [32].

**Figure 1.**
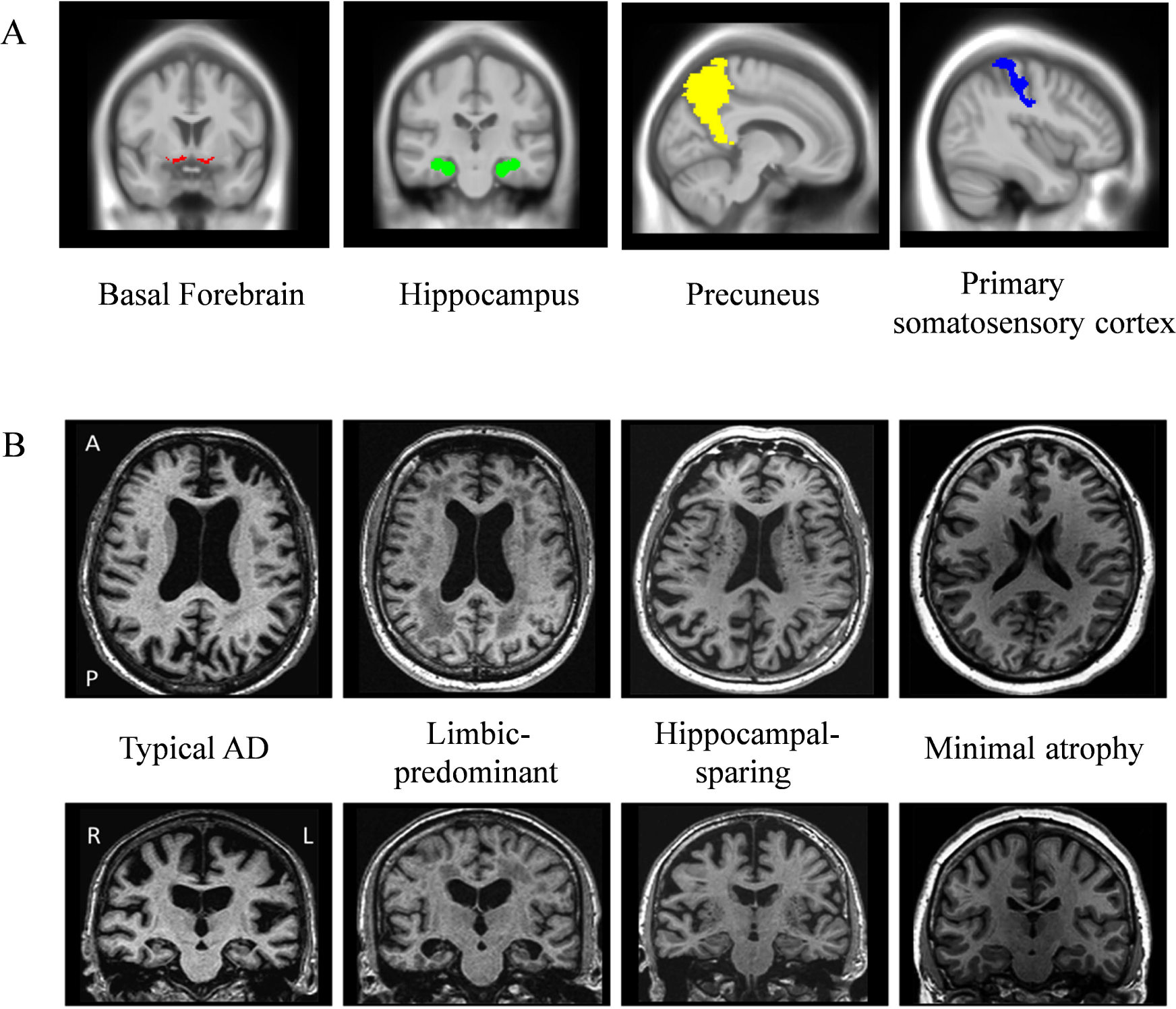
(A) Regions of interest (ROIs) depicted in colors and, (B) examples of Alzheimer’s disease subtypes. Alzheimer’s disease subtypes are based on patterns of brain atrophy classified according to the different visual rating scales. Abbreviations: A, anterior; P, posterior; R, right; L, left

#### 2.2.3. AD subtypes based on patterns of brain atrophy

All scans were rated by an experienced neuroradiologist who was blinded to participant’s information and has demonstrated excellent intra- and inter-rater reliability in patients from the ADNI cohort [6,33]. Three visual rating scales were used for subtyping as detailed elsewhere [34]. Briefly, atrophy in the medial temporal lobe was evaluated with the medial temporal atrophy (MTA) scale [35]; atrophy in the posterior cortex was evaluated with the posterior atrophy (PA) scale [36]; and atrophy in the frontal lobe was evaluated with the global cortical atrophy scale – frontal subscale (GCA-F) [33]. AD subtyping was determined by combining the scores from MTA, GCA-F, and PA, as previously described [6]. Please see Appendix A for detailed information.

### 2.4. Other measures

We selected age, sex, and years of education for the demographic description of the cohorts. Clinical variables included the clinical dementia rating (CDR) total score [37] for disease severity (very mild (0.5) and mild (1) dementia), and the mini-mental state examination (MMSE) total score [25] for global cognition. We also included APOE genotype, with presence of at least one ε4 allele considered for carriership.

### 2.5. Statistical analysis

One-way ANOVA was used for continuous and dummy variables. Spearman correlations were used to investigate the association of volume of the basal forebrain with the other ROIs. Linear mixed effect models were applied to investigate the interaction between study group (between-subjects factor, 5 levels including healthy controls and the four AD subtypes) and time (within-subjects factor, 4 levels) separately for the four brain ROIs. Estimates of volumetric change over time (mm3 per time unit) from the linear mixed effect models are reported as a measure of atrophy rate. P-values in all post-hoc analyses were adjusted using the Benjamini-Hochberg correction for multiple comparisons. Results were deemed significant when p≤.05.

## 3. Results

The AD subtypes in the ADNI cohort did not differ from each other in key clinical measures (Table 1). The NGF AD patients displayed younger age, less years of education, and lower MMSE score compared with the ADNI AD patients (Table 1).

### 3.1. Basal forebrain atrophy across AD subtypes (ADNI cohort)

At baseline, the four AD subtypes had comparable volumes of the basal forebrain (p>.05), but the volumes were reduced compared with the healthy controls (all p<.05 when uncorrected; only typical and limbic-predominant AD p<.05 when corrected for multiple comparisons) (Table S1). In contrast, longitudinal basal forebrain atrophy rates differed between the AD subtypes. The mixed model showed a significant interaction between study group (all the AD subtypes and healthy controls) and time (F_(4, 383)_= 2.407; p=.049, Figure 2A). We found a significantly faster atrophy rate in limbic-predominant AD (estimate: -17.7) compared with healthy controls (estimate: -8.1; t_(379)_=2.914, p=.004), typical AD (estimate: -10.7; t_(382)_=1.990, p=.047), and minimal atrophy AD (estimate: -7.0; t_(385)_=2.015, p=.045). No other significant differences were found in atrophy rates. All study groups except for minimal atrophy AD showed significant volume decline over time (p<.001). All the models were controlled for age, sex, and TIV (see model’s full details in Table S2).

**Figure 2.**
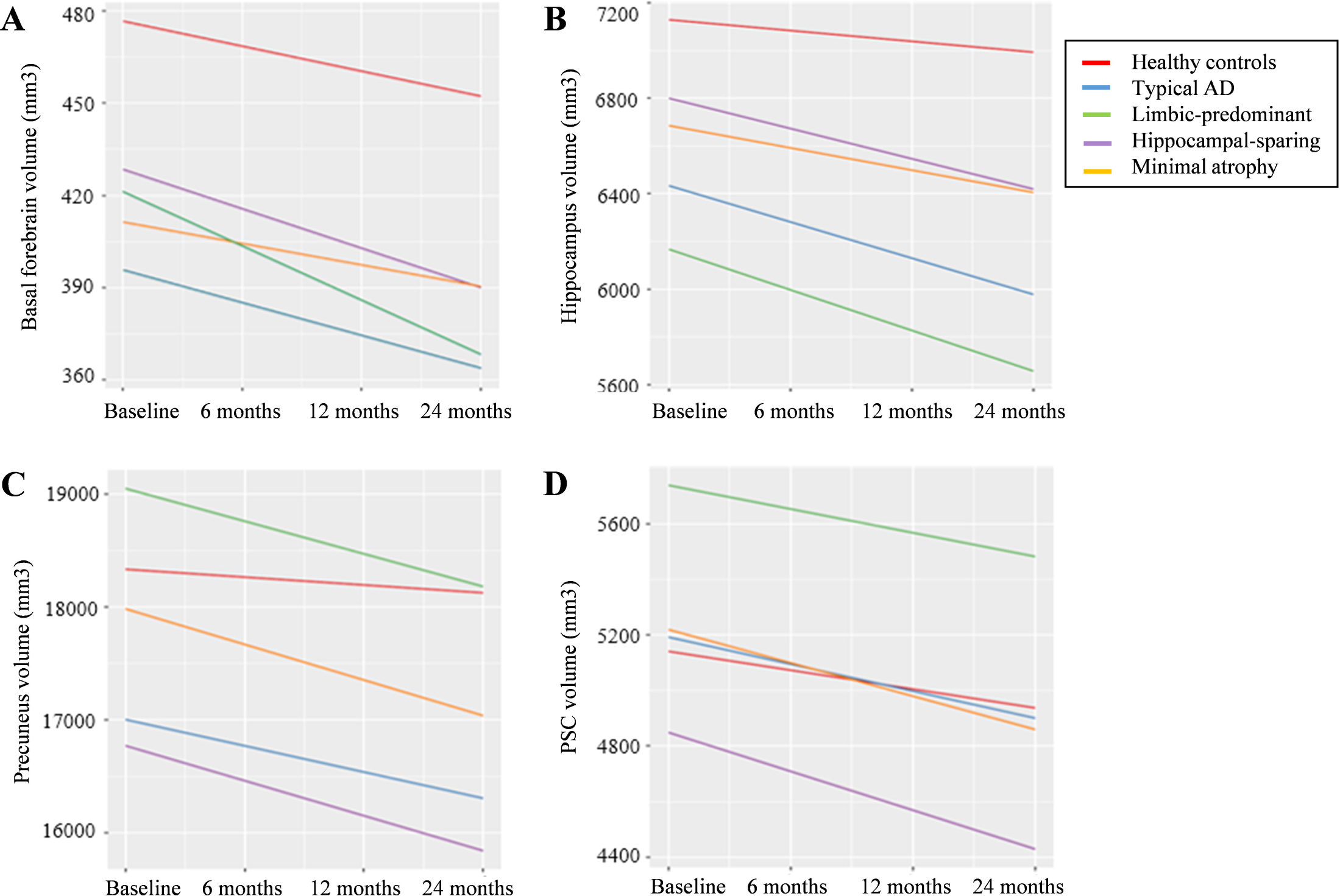
Longitudinal rates of volume change in the AD subtypes (ADNI cohort). Longitudinal rates of volume change of the (A) basal forebrain, (B) Hippocampus, (C) Precuneus, and (D) Primary somatosensory cortex (PSC) over 24 months are presented for all five studied groups. Volume estimations in the Y-axis are adjusted for age, sex and total intracraneal volume.

### 3.2. Hippocampus, precuneus, and PSC across AD subtypes (ADNI cohort)

At baseline, the AD subtypes displayed regional atrophy coherent with the definition of the AD subtypes (Table S1). The mixed model for the hippocampus showed a significant interaction between study group and time (F_(4, 378)_= 18.262; p<.001, Figure 2B). The rate of hippocampal atrophy significantly exceeded that of the healthy controls (estimate: -45.2) in typical (estimate: -151.9; t_(378)_=-7.054, p<.001), limbic-predominant (estimate: -169.9; t_(377)_=-6.419, p<.001), and hippocampal-sparing AD (estimate: -127.0; t_(378)_=-3.710, p<.001). Minimal atrophy AD (estimate: -93.6) showed a slower hippocampal atrophy rate compared to typical (t_(380)_=-1.998, p=.047) and limbic-predominant AD (t_(379)_=-2.412, p=.016). The mixed model for the precuneus showed a significant interaction between study group and time (F_(4, 361)_= 4.882; p<.001, Figure 2C). All the AD subtypes showed a faster atrophy rate than the healthy controls (p<.05), but no significant differences were found among the AD subtypes (p>.05). The mixed model for the PSC showed no significant interaction between study group and time (F_(4, 376)_= 1.218; p=.303, Figure 2D). All the models were controlled for age, sex, and TIV (see models’ full details in Table S3).

We also investigated the correlation between longitudinal atrophy of the basal forebrain (over 24 months) and longitudinal atrophy of its target regions, separately for each subtype. We obtained a moderate to weak correlation indicating that faster atrophy in the basal forebrain was associated with faster atrophy in the hippocampus in the typical AD subtype (rs= 0.393, p=.021, Figure 3). No other significant correlations were found.

**Figure 3.**
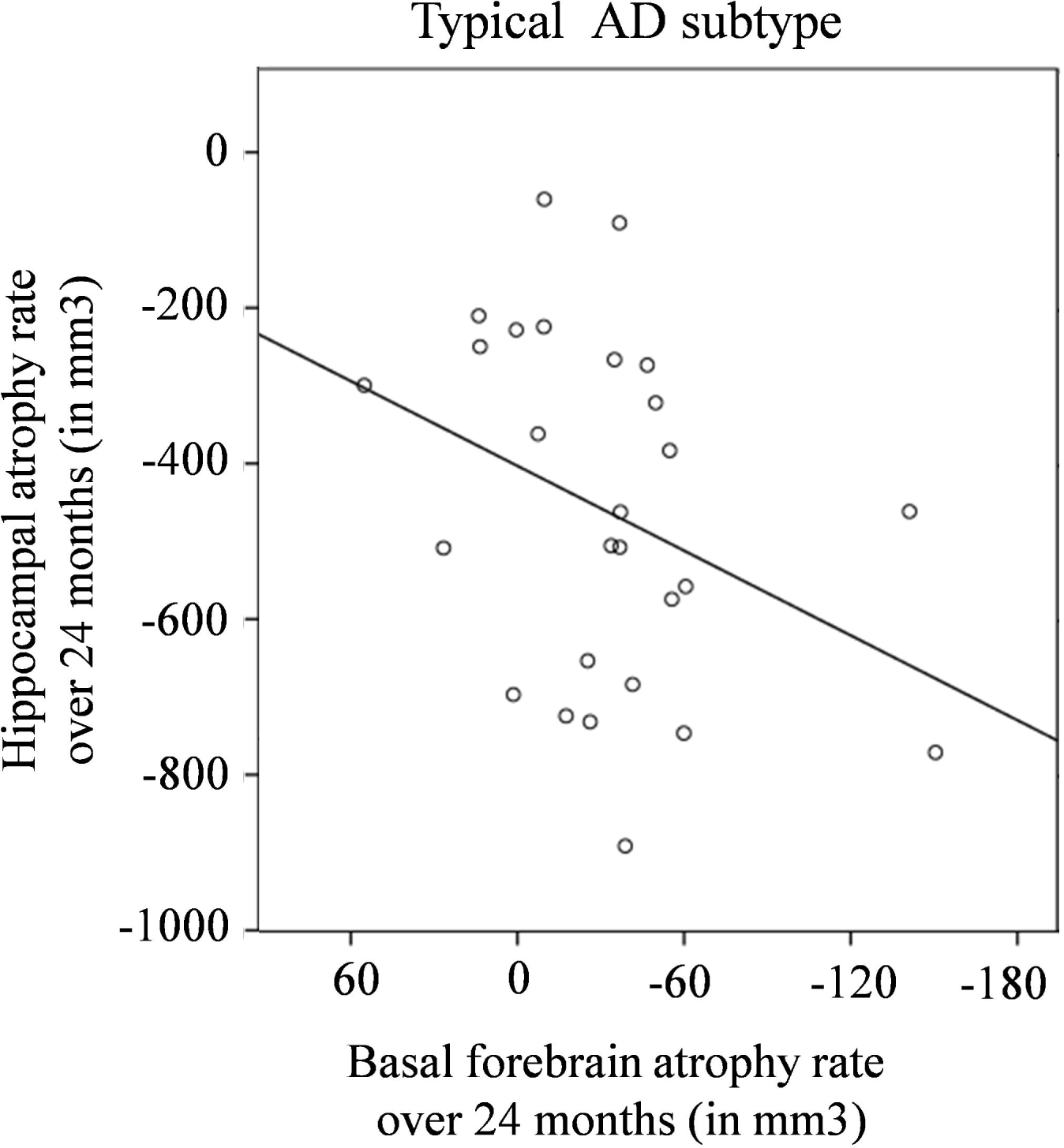
Association between longitudinal atrophy rates of the basal forebrain and the hippocampus (ADNI cohort). Longitudinal atrophy rate is calculated as volume at 24-months follow-up minus volume at baseline

### 3.3. Regional atrophy in the NGF AD patients as compared with subtype-specific ADNI data

The NGF treatment modifies the trajectories of global brain atrophy and CSF biomarkers [26,38]. In order to understand whether this effect could depend on the AD subtype, we investigated longitudinal atrophy rates of the basal forebrain and its target regions in the NGF patients by using subtype-specific cut-offs from the ADNI cohort (see details in the footnote of Figure 4) [39]. Consequently, a graphical comparison of longitudinal atrophy in AD subtypes with and without NGF treatment could be explored at 6- and 12-months follow-up (data available for the NGF cohort). Patients above the cut-off reflect slower atrophy rate than expected, hence suggesting a possible treatment effect. Figure 4 shows that NGF patients with hippocampal-sparing subtype showed slower atrophy rate of the precuneus (values above the cut-off), and NGF patients with typical and hippocampal-sparing subtypes showed slower atrophy of the hippocampus. A patient with hippocampal-sparing AD also showed slower atrophy of the PSC. Some interesting clinical observations are that the youngest patients and those with highest education, a commonly used proxy for cognitive reserve, showed the slowest hippocampal atrophy rate. Contrarily, the patient with fastest atrophy of the basal forebrain had lowest education.

**Figure 4.**
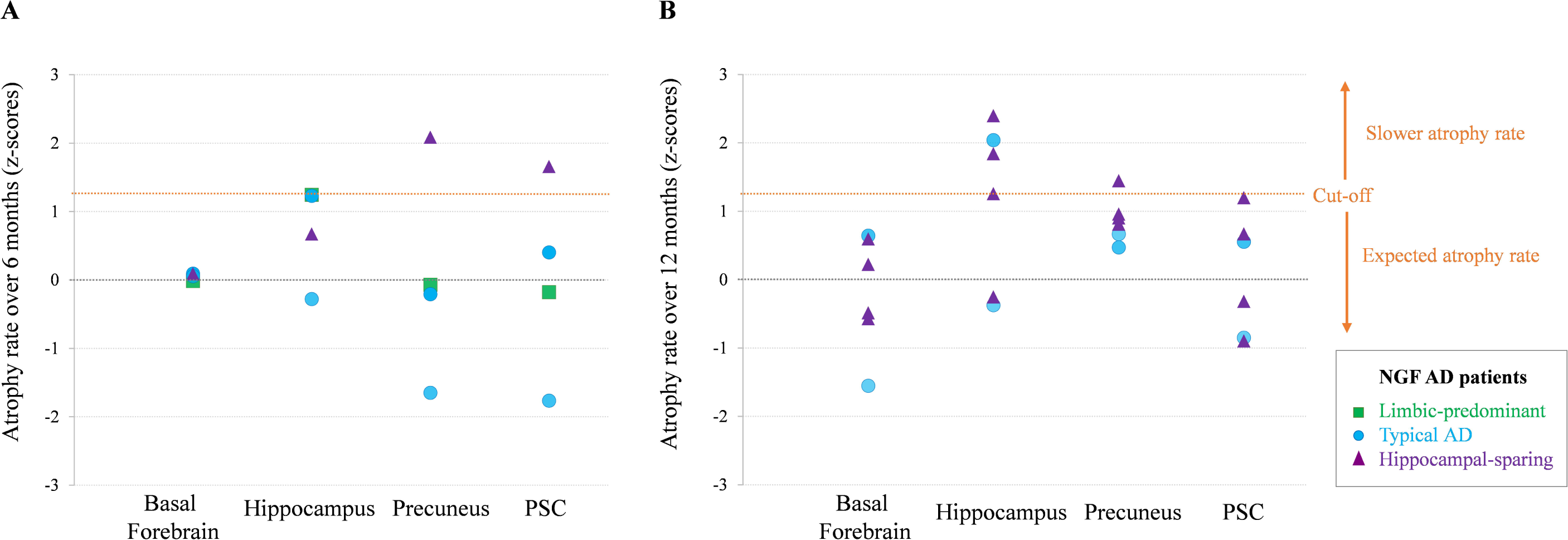
Longitudinal atrophy rates of NGF AD patients plotted against subtype-specific z-scores from the ADNI data. First, using ADNI data, we calculated subtype-specific cut-offs for longitudinal atrophy rates based on the upper 10^th^ percentile (+1.3 standard deviations) [39]. Second, NGF AD patients were classified into one of the four subtypes and their longitudinal atrophy rates were compared to the subtype-specific cut-offs derived from the ADNI data. Patients above the cut-off reflect slower atrophy rate than expected, hence suggesting a possible treatment effect. Figures A and B represent the longitudinal atrophy rates at 6 months (calculated as volume at 6-months follow-up minus volume at baseline, and transformed to z-scores using subtype-specific ADNI reference data), and at 12 months (volume at 12-months follow-up minus volume at baseline, and transformed to z-scores using subtype-specific ADNI reference data) for the basal forebrain, hippocampus, precuneus, and primary somatosensory cortex (PSC). Analyses were done separately over 6 and 12 months because MRI scans were available only at 6-month follow-up for four of the NGF patients, while they were available only at 12-month follow-up for the other six NGF patients. “Expected atrophy rate” reflects the average atrophy rate of each subtype from the ADNI cohort (Z-score = 0) with a two-tailed confidence interval of 1.3 as determined by the upper 10th percentile cut-off [39], represented by the orange dashed line. “Slower rate atrophy” represents the area from this cut-off and above, where some NGF AD patients had presumable less atrophy over time. Symbols correspond to NGF individuals’ atrophy rate. Color correspondence represents the limbic-predominant subtype (in green), typical AD subtype (in blue), and hippocampal-sparing (in purple).

## 4. Discussion

The aim of this study was to investigate (1) differences in basal forebrain volume and longitudinal atrophy rates between different subtypes of AD and healthy controls, (2) differences in longitudinal atrophy rates of the basal forebrain target regions (hippocampus and precuneus), (3) the association between basal forebrain atrophy and its target regions, and (4) regional atrophy rates of AD patients with and without a cholinergic treatment consisting of encapsulated NGF biodelivery to the basal forebrain. All four AD subtypes showed comparable volume of the basal forebrain at baseline, while they all showed significantly reduced volume of the basal forebrain compared with the healthy controls. Further, limbic-predominant AD showed faster basal forebrain atrophy over 24 months as compared with the other subtypes, whereas the basal forebrain volume in minimal atrophy AD did not change significantly over time. Compared to healthy controls, all AD subtypes showed faster atrophy in the hippocampus (except for the minimal atrophy subtype) and precuneus, but not in the PSC. Further, the basal forebrain atrophy rate was significantly associated with hippocampal atrophy rate in typical AD. The NGF treatment seemed to slow down atrophy rates of the precuneus and hippocampus, but not of the basal forebrain. Importantly, this effect was largely dependent on the AD subtype, suggesting the best response in hippocampal-sparing AD patients.

The study of AD subtypes has attracted great attention in the last years. Despite an increasing number of publications, possible differences in cholinergic system integrity across subtypes have not yet been investigated systematically. In a voxel-wise analysis, Dong et al. (2016) reported clusters of reduced gray matter volume corresponding to the basal forebrain in two AD subtypes that resembled limbic-predominant and typical AD. Hence, our study is to our knowledge the first systematic investigation of the basal forebrain across established atrophy subtypes of AD, including both cross-sectional and longitudinal data. Extending the incidental findings in the voxel-wise study of Dong et al. (2016), we demonstrated that cholinergic basal forebrain volume is reduced in all AD subtypes, including minimal atrophy AD. This is a relevant finding raising the hypothesis that cholinergic disruption due to neurodegeneration of the basal forebrain may be the basis of the clinical symptoms in the absence of overt cortical atrophy in minimal atrophy AD. Similar findings in pre-dementia stages support this hypothesis. For instance, volume of the basal forebrain was found to be atrophied and to correlate with reduced cognition in pre-dementia patients lacking overt cortical atrophy [30,41]. Further, atrophy in the basal forebrain appears to precede atrophy in the medial temporal lobe [31,42] in the development of AD. The pathophysiological explanation that possibly underlies this finding is that the basal forebrain is among the earliest sites for pre-tangle lesions [43].

Even though all AD subtypes displayed comparable basal forebrain volume at baseline, the longitudinal atrophy rate of the basal forebrain was different. Limbic-predominant AD showed fastest progression and minimal atrophy AD showed no significant decline over time, with typical and hippocampal-sparing AD having an intermediate atrophy rate. Limbic-predominant AD in the ADNI-1 cohort is among the subtypes displaying fastest cognitive decline [6]. Also, AD patients with high atrophy in the medial temporal lobe are known to respond worse to ChEI [13], which could be related to the fast neurodegeneration rate of the basal forebrain in this subtype. These findings highlight a possible inter-relation between atrophy of the basal forebrain, patterns of cortico-limbic atrophy, disease progression, and, possibly, treatment response. Baseline volumes and longitudinal atrophy of the hippocampus and precuneus were as expected and coherent with the definition of the AD subtypes (e.g. larger hippocampal atrophy in typical and limbic-predominant AD; larger precuneus atrophy in hippocampal-sparing and typical AD; etc.). To our knowledge, no previous subtype studies have reported longitudinal atrophy rates for these two brain regions.

The discussion above highlights the role of the basal forebrain as part of a large network projecting to the hippocampus and diffuse neocortical association regions [12,44]. Differential involvement of this cholinergic network could be the basis of different patterns of neurodegeneration in these AD subtypes. Limbic-predominant atrophy would stem from disruption of basal forebrain projections to the hippocampus, whereas hippocampal-sparing atrophy may be related to non-hippocampal projections to regions such as the precuneus. Typical AD would include disruption of both types of projections. We attempted to test this hypothesis by investigating subtype-specific explorative correlations between the atrophy rate of the basal forebrain and atrophy rates of the hippocampus and precuneus, respectively. We only found a significant association with the hippocampus in typical AD. A similar association has previously been found in cross-sectional analyses of cohorts including a heterogeneous group of AD patients [30,41]. Although we did not find a correlation between atrophy of the basal forebrain and atrophy of the precuneus in hippocampal-sparing and typical AD, such a correlation has been found in cross-sectional studies of heterogeneous AD cohorts as well [30,41,45]. Future studies including connectivity analyses in larger subtype groups may be of relevance for testing this hypothesis further. For example, we recently found that fronto-parietal and occipital networks are altered in both typical and hippocampal-sparing AD, but not in limbic-predominant AD [46].

Considering the potential relevance of these subtypes for precision medicine interventions, we explored subtype-specific effects of NGF treatment on rates of regional atrophy in 10 patients. While this NGF cohort from a phase 1 study is too small for valid statistical testing, this is the first MRI study exploring the potential relevance of AD subtypes for detecting effects of cholinergic treatment. Our findings suggest different response to the NGF treatment depending on the AD subtype. Among the three subtypes available in the NGF cohort (i.e. typical, limbic-predominant, and hippocampal-sparing AD), hippocampal-sparing AD seemed to have the best response to the NGF treatment. A possible explanation for this finding is that AD patients with high atrophy in the medial temporal lobe are known to respond less to pharmacologic cholinergic treatment [13], whereas hippocampal-sparing AD lacks atrophy in the medial temporal lobes. Another potential explanation is comorbidity with dementia with Lewy bodies (DLB)-related pathology in this subtype. Hippocampal-sparing atrophy is the most frequent pattern of atrophy in DLB patients [14], who also respond better to cholinergic treatment than AD patients do [15]. We observed that younger age and higher cognitive reserve may also positively influence treatment response, in agreement with the conclusions from our previous study on the NGF cohort [26].

One limitation of this study is the small sample size of the NGF cohort. We thus consider the NGF part as exploratory, and we report those results as preliminary but of interest due to the unique nature of that dataset. In addition, longitudinal data for the hippocampal-sparing and minimal atrophy AD subtypes at 12- and 24-month follow-up in the ADNI-1 cohort was limited. We used mixed effect models, which are superior on small groups and censored longitudinal data [47]. Future studies including larger cohorts are warranted. Further, meaningful translational approaches, such as crossing validation of the subtype-stratification with the genetic sequencing could be considered. Our interpretations on treatment effects are based on the NGF treatment, which is an add-on treatment since all the patients were on stable ChEI treatment before and during the study. It is possible that the effects reported here are even larger in drug naïve AD patients, but this is difficult to test because patients in most of the available AD cohorts are under symptomatic treatment. Moreover, one must consider the effect of the combined administration of NGF and cholinesterase inhibitors, as these two agents can modulate several signaling pathways that are known to affect neurogenesis, synaptic modulation, and regeneration [48–50].

Finally, connectivity analyses using other imaging modalities such as diffusion tensor imaging or functional MRI at the resting state [51] might shed further light on how the cholinergic system contributes to treatment response in the different AD subtypes. Unfortunately, we did not have these data available on the NGF cohort, and only a subset of the ADNI-1 cohort includes these data.

## 5. Conclusions

The heterogeneity within AD is one of the greatest challenges for the development of successful disease modifying drugs. Precision medicine based on disease biomarkers has recently emerged as one of the most promising strategies to guide AD research, drug discovery, and clinical disease management. Distinct atrophy subtypes of AD are now well recognized but there is still a long way to completely understand the mechanisms and modulating factors underlying these subtypes. Such understanding is needed because these mechanisms and modulating factors may determine differential treatment response across subtypes. This is a very attractive field and approach but very few data exist yet. Our current study is the first in exploring differences in cholinergic system degeneration across different subtypes of AD and further points to the possible relevance of these differences in predicting response to cholinergic treatment.

## Data Availability

The ADNI dataset used and analyzed during the current study are available from the corresponding author on reasonable request. The NGF dataset generated and/or analyzed are not publicly available due to compromise of individual privacy of these patients but are available from the corresponding author on reasonable request.

## 6. Abbreviations

*A β*: Amyloid β
*AD*: Alzheimer’s disease
*ADNI*: Alzheimer’s Diseases Neuroimaging Initiative
*ANOVA*: Analysis of variance
*APOE*: Apolipoprotein E
*CDR*: Clinical dementia rating
*ChEI*: Cholinesterase inhibitors
*CSF*: Cerebrospinal fluid
*DARTEL*: Diffeomorphic anatomical registration through exponentiated lie algebra
*DLB*: Dementia with Lewy bodies
*ECB*: Encapsulated cell biodelivery
*GCA-F*: Global cortical atrophy – Frontal
*GM*: Gray matter
*MMSE*: Mini-Mental State Examination
*MNI*: Montreal Neurological Institute
*MRI*: Magnetic resonance imaging
*MTA*: Medial temporal atrophy
*NFT*: Neurofibrillary tangle
*NGF*: Nerve growth factor
*NINCDS-ADRDA*: National Institute of Neurological and Communicative Disorders and Stroke and the Alzheimer’s Disease and Related Disorders Association
*PA*: Posterior atrophy
*PSC*: Primary somatosensory cortex
*ROI*: Region of interest
*SPM*: Statistical parametric mapping
*TIV*: Total intracranial volume
*VBM*: Voxel-based morphometry
*WM*: White matter

## Declarations

### Ethics approval and consent to participate

The study protocol and informed consent form were reviewed and approved by the institutional review board of each center. For the NGF, the Swedish Medical Products Agency and the Regional Human Ethics Committee of Stockholm approved the study (Dnr 2007/986-31/3). The patients and their caregivers gave us written informed consent prior to study inclusion.

### Consent for publication

Informed consent for the publication of data was obtained from the participants or their caregivers.

### Competing interests

Lars Wahlberg is an owner and employee of Gloriana Therapeutics, which developed the NGF secreting device. All the other authors declared no conflicts of interest.

### Funding

This project is financially supported by the Swedish Foundation for Strategic Research (SSF), the Swedish Research council (VR, grant #2016-02317), the regional agreement on medical training and clinical research (ALF) between Stockholm County Council and Karolinska Institutet, the Strategic Research Programme in Neuroscience at Karolinska Institutet (StratNeuro), the Swedish Alzheimer foundation, the Swedish Brain foundation, Åke Wiberg foundation, Åhlen foundation, King Gustaf V’s and Queen Victoria’s Foundation of Freemason, Olle Engkvist Byggmästare foundation, Ragnhild and Einar Lundström Minne, Gun and Bertil Stohnes, Sigurd och Elsa Goljes Minne, Ålderssjukdomar, Gamla Tjänarinnor, Karolinska Institutet Forskningstiftelse, Demensförbundet. We also thank Birgitta and Sten Westerberg for additional financial support.

Data collection and sharing for the ADNI study was funded by the Alzheimer’s Disease Neuroimaging Initiative (ADNI, National Institutes of Health Grant U01 AG024904) and DOD ADNI (Department of Defense award number W81XWH-12-2-0012). ADNI is funded by the National Institute on Aging, the National Institute of Biomedical Imaging and Bioengineering, and through generous contributions from the following: Alzheimer’s Association; Alzheimer’s Drug Discovery Foundation; BioClinica, Inc.; Biogen Idec Inc.; Bristol-Myers Squibb Company; Eisai Inc.; Elan Pharmaceuticals, Inc.; Eli Lilly and Company; F. Hoffmann-La Roche Ltd and its affiliated company Genentech, Inc.; GE Health-care; Innogenetics, N.V.; IXICO Ltd.; Janssen Alzheimer Immuno-therapy Research & Development, LLC.; Johnson & Johnson Pharmaceutical Research & Development LLC.; Medpace, Inc.; Merck & Co., Inc.; Meso Scale Diagnostics, LLC.; NeuroRx Research; Novartis Pharmaceuticals Corporation; Pfizer Inc.; Piramal Imaging; Servier; Synarc Inc.; and Takeda Pharmaceutical Company. The Canadian Institutes of Health Research provides funds to support ADNI clinical sites in Canada. Private sector contributions are facilitated by the Foundation for the National Institutes of Health (www.fnih.org). The grantee organization is the Northern California Institute for Research and Education, and the study is coordinated by the Alzheimer’s disease Cooperative Study at the University of California, San Diego. ADNI data are disseminated by the Laboratory for Neuro Imaging at the University of California, Los Angeles.

### Authors’ contributions

AM and DF carried out the analyses and wrote the manuscript. MG contributed to the structural longitudinal analysis. DF, EW, AM and ME conceived and designed the study. AM prepared the figures and tables. All authors reviewed and accepted the current version of the manuscript.

Data used in preparation of this article were obtained from ADNI database (adni.loni.usc.edu). Investigators within the ADNI contributed to the design and implementation of ADNI and/or provided data but did not participate in analysis or writing of this report. A complete listing of ADNI investigators can be found at: http://adni.loni.usc.edu/wp-content/uploads/how_to_apply/ADNI_Acknowledgement_List.pdf

## Acknowledgements

Not applicable

## Notes

### Clinical Trial

NCT01163825

